# Socioeconomic deprivation and time trends in pediatric hospital admissions, intensive care treatment, and mortality: a nationwide population-based study in Germany

**DOI:** 10.64898/2026.08.15.26360488

**Authors:** Rayan Hojeij, Carina Oenning, Habyna Ravichandrajah, Christoph Härtel, Christian Dohna-Schwake, Ursula Felderhoff-Müser, Nora Bruns

## Abstract

**Background:** Socioeconomic deprivation is associated with childhood morbidity, but nationwide evidence on critical illness and death in a health system with universal insurance coverage is scarce. We assessed the association between area-level deprivation and the population-level incidence of hospital admission, complex intensive care treatment (CICT), and CICT-related mortality in German children, and changes over time.

**Methods:** Population-based analysis of complete German hospital discharge data, 2016 to 2023, covering all cases aged > 28 days to < 18 years. Cases were linked to the German Index of Socioeconomic Deprivation (GISD) via the municipality of residence and grouped into quintiles (Q1 least, Q5 most deprived). Incidence rates were calculated per 100,000 child years. Negative binomial regression adjusted for calendar year, with population as offset, yielded adjusted incidence rate ratios (aIRR) per one-quintile increase in deprivation; sensitivity analyses additionally adjusted for age group. Excess cases were estimated by applying Q1 incidence rates to Q2 to Q5.

**Results:** Of 8,890,103 pediatric cases, 140,509 (1.6 %) received CICT and 3,386 (2.40 %) of these died. Incidence rose with deprivation from Q1 to Q5: admissions 6,191 to 9,255 per 100,000 child years, CICT 97 to 128, mortality 2.54 to 2.96. Each one-quintile increase was associated with higher risk of admission (aIRR 1.10, 95 % CI 1.10-1.11), CICT (1.07, 1.05-1.08), and mortality (1.04, 1.01-1.06); estimates were unchanged after age adjustment. Relative to Q1 rates, Q2 to Q5 accounted for 1,295,896 excess admissions (20.8 %), 11,254 excess CICT cases (12.6 %), and 194 excess deaths (8.7 %). Case fatality among CICT cases was lower in more deprived quintiles (2.35 % in Q5 versus 2.64 % in Q1), as were organ dysfunction and chronic conditions. Disparities in admission and CICT narrowed over time, whereas the mortality gradient persisted.

**Conclusions:** Universal health insurance did not eliminate socioeconomic inequalities in pediatric critical illness. Deprivation increased the population burden of admission, intensive care, and death, but did not worsen outcomes once intensive care had begun, indicating that inequalities arise before pediatric intensive care and that prevention upstream in the care continuum is the primary target.

## Introduction

Social deprivation is a well-established risk factor for health inequalities in children. A recent multicenter, retrospective study highlighted the interaction between neighborhood-level deprivation and worsened health disparities in hospitalized children [1]. Children living in deprived districts in Glasgow were about nine times more likely to be admitted to hospital for any reason than children in non-deprived districts [2], and deprivation has been linked to multiple admissions for organic reasons, driven by both organic pathology and impaired parental coping [3]. This gradient extends to neonatal intensive care, where social deprivation correlates with neonatal morbidity, neonatal unit admission, and mortality [4–6].

In pediatric intensive care, US children with greater socioeconomic vulnerability had increased odds of pediatric intensive care unit (PICU) mortality, with Black children at particularly increased risk of both PICU admission and mortality [7]. Variation in PICU mortality by ethnicity and area-level deprivation has similarly been observed in the UK [8, 9], and a clear gradient of increasing child mortality has been described across England as deprivation increases, with little variation by area, age, or other demographic factors [10].

However, existing evidence stems predominantly from North America and the United Kingdom (UK), with mainland Europe being underrepresented. Many studies are based on regional analyses, while findings may not be directly transferable between healthcare systems. In the US, for example, the prevalence of children receiving ICU care has increased over time [11], contrasting the ongoing reduction of pediatric inpatient capacity and admissions in Germany [12–14]. The German hospital dataset (GHD), a nationwide diagnosis related groups (DRG) hospital discharge statistics, encompasses a complete census of all acute-care hospital admissions and can be linked to a validated regional deprivation index (GISD) [15], thereby providing a unique opportunity to investigate how socioeconomic deprivation influences the population burden of pediatric critical illness requiring intensive care.

The aim of this study was to assess the association between area-level socioeconomic deprivation and the population-level incidence of hospital admission, complex intensive care treatment (CICT) according to operation and procedure codes (OPS), and in-hospital mortality among children in Germany, and to evaluate whether these associations changed over time between 2016 and 2023. CICT incidence was defined *a priori* as the primary outcome, with hospital admission and in-hospital mortality as secondary outcomes.

## Methods

This retrospective observational study used nationwide routine health care data from the German hospital dataset (GHD). Cases were identified via age at admission and analyzed with respect to performed complex intensive care treatment. The study included the datasets from 2016 - 2023 [16–23] that were matched with the German Index of Socioeconomic Deprivation based on the area key of the place of residence of each case.

### Data source

The GHD is a nationwide dataset that contains all discharges from public hospitals in Germany. Because there are no private children’s hospitals or private pediatric intensive care units in Germany, the dataset represents a complete nationwide census of pediatric hospital admissions. Since 2004, German hospitals receive compensation based on Diagnosis Related Groups (DRG). As per §21 KHEntgG, it is mandated by law that German hospitals share data on all hospital discharges with the Hospital Remuneration Institute (InEK). After passing plausibility checks, anonymized data is forwarded to the Federal Statistical Office (FSO). Upon request, scientists can gain access to this dataset or subsets at regional research data centers.

### Case selection

Cases > 28 days (age at admission) and < 18 years of age discharged from a German hospital between 2016 and 2023 were analyzed. Cases were classified into ICU and non-ICU cases based on OPS that indicate CICT.

### Complex intensive care treatment

The code for pediatric complex intensive care treatment (CICT) can be applied if certain criteria are fulfilled while a patient is treated in an intensive care unit (Supplementary table 1). The code for adult CICT requires similar prerequisites and can be applied from 14 years onwards. The pediatric CICT cannot be applied in neonates (age at admission < 28 days) including preterm infants during their primary stay in hospital, who were therefore excluded. For adolescents ≥ 14 years, either the code for pediatric or adult CICT could be applied in the first part of the study, but since 2021 only certified pediatric intensive care units can apply the pediatric CICT after structural certification (according to *Strukturprüfungs-Richtlinie*, StrOPS-RL), with a transition phase throughout 2021 and 2022. The analyzed dataset covers the transition of CICT coding, causing mixing of pediatric and adult CICT codes and new coding prerequisites.

### Determination of socioeconomic deprivation

The German Index of Socioeconomic Deprivation (GISD) is a validated, area-level composite index developed by the Robert Koch Institute to quantify regional socioeconomic deprivation in Germany [15, 24]. It is based on municipal or district mappings and combines indicators of education, employment, and income into a standardized deprivation score. It is widely used to assess health equity when individual socioeconomic data are unavailable. GISD scores are available for different administrative levels and are commonly categorized into quintiles ranging from the least deprived (Q1) to the most deprived (Q5). The GISD was assigned at the municipality level using the official municipality code (*Amtlicher Gemeindeschlüssel*, AGS).

### Population at risk

Nationwide annual pediatric population denominators were calculated specifically for each GISD quintile. Municipality-level population data were obtained from the German Federal Institute for Research on Building, Urban Affairs and Spatial Development (*Indikatoren und Karten zur Raum- und Stadtentwicklung*, www.inkar.de, accessed May 5, 2026). Municipality-level population data were linked to the German Index of Socioeconomic Deprivation (GISD) using the AGS.

### Organ dysfunction

The Pediatric Organ Dysfunction Index (PODI) is a pragmatic severity-adjustment tool derived from ICD-10 diagnosis codes and German Operation and Procedure System (OPS) codes to approximate acute organ dysfunction using administrative healthcare data. It mirrors the organ-system domains of the pediatric Sequential Organ Failure Assessment (pSOFA) score [25].

### Chronic conditions

Medical complexity arising from chronic conditions was determined using the second version of the Pediatric Complex Chronic Conditions (PCCC) system [26]. We applied the previously described method with the same minor modifications necessitated by the lack of information on device prescription in the GHD [12].

### Primary and secondary outcomes

The unit of analysis was a pediatric case discharged from a German hospital between 2016 and 2023. The primary outcome was the population-level incidence rate (IR) of CICT per GISD quintile during the study period. Secondary outcomes included IRs for hospital admissions and in-hospital mortality of CICT cases.

### Missing data

#### Hospital discharge data (GHD)

There were no missing data on age, and all cases had at least one diagnosis code and one department code. Missing OPS codes could not be identified, as it is not possible to distinguish between procedures that were not performed and those that were not coded. However, because procedural codes and relevant secondary diagnoses directly affect DRG reimbursement, systematic undercoding is considered unlikely.

#### Population denominators

There were no missing data for any municipality or year.

#### GISD assignment

Because GISD data were available only through 2021 at the time of analysis, the 2021 classification was applied from 2022 and 2023.

- <u>Missing AGS in the GHD:</u> Cases with no AGS available were assumed to have their place of residency outside of Germany or – for German residents – would be missing at random and were excluded from all further analyses.
- <u>Linkage failure in population data and GHD:</u> No GISD classification is available for 204 municipality-free administrative areas. Cases from these areas were excluded from GISD-stratified analyses. Because most unmatched cases in the GHD originated from Berlin, population denominators were adjusted accordingly. As a sensitivity analysis, all Berlin cases were alternatively assigned to Berlin’s GISD quintile (AGS 11000000)(years 2016 – 2018: 4^th^ quintile, years 2019 – 2023: 3^rd^ quintile).

Due to statutory cell-size censoring requirements, a small number of deaths (n = 43) from cases admitted in 2015 but discharged in 2016 could not be excluded from the age-adjusted mortality models, unlike in the calendar-year-stratified analyses, resulting in a minor difference in total death counts between the two approaches.

### Statistical Analyses

Categorical variables are presented as counts and percentages. Continuous variables are reported as mean ± standard deviation (SD) or median with interquartile range (IQR), as appropriate. Descriptive analyses were performed overall and stratified by GISD quintile.

Incidence rates (IRs) of hospital admission, CICT, and in-hospital mortality of CICT cases were calculated using the corresponding annual pediatric population overall or within each GISD quintile as the denominator and are reported per 100,000 child years (CY). Annual IRs were additionally calculated overall and for each GISD quintile to assess temporal trends. Standardized incidence ratios (SIRs) with 95% confidence intervals (CIs) were calculated for each study year using the pooled incidence rate across the entire study period as the reference.

The association between socioeconomic deprivation (GISD quintile) and each outcome was quantified using negative binomial regression, adjusted for calendar year, with population as offset. Model selection was based on the test results for overdispersion using a likelihood-ratio-test (LRT). Results are reported as adjusted incidence rate ratios (aIRR) per one-quintile increase in GISD, with 95% confidence intervals (CI). Time-trend heterogeneity was assessed by comparing models with and without a quintile-by-year interaction term using a likelihood-ratio test.

To estimate the burden attributable to the observed socioeconomic gradient, the pooled incidence rate in the least deprived quintile (Q1) over the study period was applied to the population of each quintile to derive expected case counts under a counterfactual scenario of uniform Q1-level risk. The difference between observed and expected cases across quintiles Q2–Q5 was summed to estimate excess cases associated with socioeconomic deprivation, reported as an absolute number and as a percentage of observed cases. This was performed separately for hospital admissions, CICT, and mortality.

Sensitivity analyses with alternative handling of unmatched Berlin cases were conducted to calculate overall IRs for hospital admission, CICT and mortality.

As a further sensitivity analysis, all IRR regression models were additionally adjusted for age group (< 3, 3 to < 6, and 6 to < 18 years). For these models, case counts and population denominators were stratified by GISD quintile, calendar year, and age group, with the age-group-specific population as offset. A likelihood-ratio test confirmed that inclusion of age group improved model fit for all three outcomes (p < 0.001); a quintile-by-age-group interaction term was tested and was not statistically significant for any outcome (hospital admission p = 0.87, CICT p = 0.67, mortality p = 0.99), so age group was retained as an additive term.

### Software

SAS Version 9.4 (SAS Institute, Cary, USA) was used to analyze and extract data at the regional research data center. RStudio using R Version 4.5.2 (Posit PBC, Boston, USA) was used for final calculations and to produce figures.

### Ethics approval

No ethics approval was required according to local legislation because we used exclusively anonymized secondary health care data and publicly available population statistics.

## Results

### Overall numbers and matching in the clinical dataset

The original GHD dataset contained 15,251,642 cases, of which 6,306,611 (41.4 %) were ≤ 28 days at admission and thus excluded (Supplementary figure 1). Another 54,923 (0.6 %) cases were excluded because of missing AGS. Out of the remaining 8,890,103 pediatric cases, 140,509 (1.6 %) received CICT and were analyzed for overall cohort analyses.

For GISD-specific analyses, linking failure produced relevant subgroups that were handled specifically: 326,717 (3.7 %) of all cases could not be matched to a GISD quintile due to incompatible AGS. 301,861 (92.4 %) of all unmatched cases originated from Berlin. Out of 14,438 unmatched CICT cases, 14,159 (98.1 %) were from Berlin. Assigning Berlin’s GISD quintiles to Berlin cases reduced unmatched cases to 24,856 (0.3 % of all cases) and 279 (1.9 % of CICT cases), respectively. Supplementary figure 1 presents the precise flow and patient number per step and analysis.

### Overall numbers and matching in the population data

Out of 123,083,920 child years (CY) throughout the study period, 6,841 could not be matched. The remaining 123,077,079 CY were unevenly distributed between the quintiles, with the highest amount in the 1^st^ quintile (n = 37,644,947) and the lowest amount in the 4 and 5 quintiles (n = 18,581,041 and n = 19,263,425) (Table 1). 52 % of CY derived from the 1 and 2 quintiles.

**Table 1:** Case characteristics of CICT cases in Germany (2016 – 2023) overall and by quintile of social deprivation.

|  | Overall | Quintile<br>1st | 2nd | 3rd | 4th | 5th |
| --- | --- | --- | --- | --- | --- | --- |
| Total pediatric end-of-year population 2016 – 2023 (child years), *n (row %)* | 123,083,920 | 37,644,947<br>(30.6 %) | 26,399,465<br>(21.4 %) | 21,188,202<br>(17.2 %) | 18,581,041<br>(15.1 %) | 19,263,425<br>(15.7 %) |
| Pediatric end-of-year population 2016 – 2023 excluding Berlin, used for main analysis (child years), *n (row %)* | 117,716,040 | 37,644,947<br>(32.0 %) | 26,399,465<br>(22.4 %) | 18,139,108<br>(15.4 %) | 16,262,254<br>(13.8 %) | 19,263,425<br>(16.4 %) |
| Pediatric hospital admissions, *n (row %)* | 8,890,103 | 2,330,495<br>(27.2 %) | 1,788,815<br>(20.9 %) | 1,350,967<br>(15.8 %) | 1,310,228<br>(15.3 %) | 1,782,881<br>(20.8 %) |
| < 3 years, *n (row %)* | 2,777,144 | 756,004<br>(28.4 %) | 555,523<br>(20.9 %) | 402,381<br>(15.1 %) | 398,420<br>(15.0 %) | 550,450<br>(20.7 %) |
| 3 to < 6 years, *n (row %)* | 1,386,576 | 370,113<br>(27.7 %) | 282,085<br>(21.1 %) | 206,018<br>(15.4 %) | 204,028<br>(15.3 %) | 274,239<br>(20.5 %) |
| 6 to < 18 years, *n (row %)* | 4,726,383 | 1,204,378<br>(26.4 %) | 951,207<br>(20.8 %) | 742,568<br>(16.3 %) | 707,780<br>(15.5 %) | 958,192<br>(21.0 %) |
| Deaths among all admissions *n (column %)* | 7,671 (0.09 %) | 2,276 (0.10 %) | 1,640 (0.09 %) | 1,160 (0.09 %) | 1,094 (0.08 %) | 1,501 (0.08 %) |
| Death within 24 hours of all deaths, *n (column %)* | 2,497 (32.6 %) | 749 (32.9 %) | 530 (32.3 %) | 358 (30.9 %) | 348 (31.8 %) | 512 (34.1 %) |
| ICU admissions, *n (row %)* | 140,509 | 36,800 (29.2 %) | 26,598 (21.1 %) | 19,044 (15.1 %) | 18,949 (15.0 %) | 24,680 (19.6 %) |
| < 3 years, *n (row %)* | 63,074 | 16,756 (29.7 %) | 11,899 (21.1 %) | 8,206 (14.5 %) | 8,420 (14.9 %) | 11,203 (19.8 %) |
| 3 to < 6 years, *n (row %)* | 19,091 | 4,968 (29.4 %) | 3,653 (21.6 %) | 2,482 (14.7 %) | 2,566 (15.2 %) | 3,247 (19.2 %) |
| 6 to < 18 years, *n (row %)* | 58,344 | 15,076 (28.6 %) | 11,046 (21.0 %) | 8,356 (15.9 %) | 7,963 (15.1 %) | 10,230 (19.4 %) |
| Sex female, *n (%)* | 61,728 (43.9 %) | 16,230 (44.1 %) | 11,606 (43.6 %) | 8,411 (44.2 %) | 8,332 (44.0 %) | 10,733 (43.5 %) |
| Case fatality in ICU cases, *n (%)* | 3,386 (2.40 %) | 973 (2.64 %) | 698 (2.62 %) | 498 (2.61 %) | 473 (2.50 %) | 579 (2.35 %) |
| < 3 years, *n (%)* | 1,567 (2.48 %) | 436 (2.60 %) | 343 (2.88 %) | 230 (2.80 %) | 229 (2.72 %) | 246 (2.20 %) |
| 3 to < 6 years, *n (%)* | 420 (2.20 %) | 130 (2.62 %) | 79 (2.16 %) | 60 (2.42 %) | 60 (2.34 %) | 78 (2.40 %) |
| 6 to < 18 years, *n (%)* | 1,399 (2.40 %) | 407 (2.70 %) | 276 (2.50 %) | 208 (2.49 %) | 184 (2.31 %) | 255 (2.49 %) |
| Survivors: |  |  |  |  |  |  |
| Mechanical ventilation, *n (%)* | 44,061 (32.1 %) | 13,355 (37.3 %) | 9,237 (35.7 %) | 6,385 (34.4 %) | 5,819 (31.5 %) | 6,934 (28.8 %) |
| Duration of mechanical | 80 (34 – 184) | 77 (32 – 175) | 76 (33 – 176) | 81 (33 – 181) | 82 (34 – 184) | 86 (35 – 192) |
| ventilation (hours), *median (IQR)* |  |  |  |  |  |  |
| LOS (days), *median (IQR)* | 9 (4 – 16) | 9 (5 – 17) | 9 (5 – 16) | 9 (5 – 16) | 9 (5 – 16) | 9 (4 – 15) |
| Deceased: |  |  |  |  |  |  |
| Mechanical ventilation, n (%) | 3,077 (92.1 %) | 902 (92.7 %) | 642 (92.0 %) | 451 (90.6 %) | 422 (89.2 %) | 517 (89.3 %) |
| Duration of mechanical ventilation (hours), *median (IQR)* | 144 (56 – 435) | 150 (57 – 437) | 153 (52 – 470) | 131 (52 – 376) | 151 (58 – 426) | 141 (59 – 416) |
| LOS (days), *median (IQR)* | 10 (3 – 33) | 10 (3 – 33) | 10 (4 – 36) | 9 (3 – 31) | 10 (4 – 31) | 9 (4 – 32) |
| Organ dysfunction |  |  |  |  |  |  |
| PODI = 0 | 86,519 (61.6 %) | 20,552 (55.9 %) | 15,362 (57.8 %) | 11,247 (59.1 %) | 11,892 (62.8 %) | 16,028 (64.94 %) |
| PODI = 1 | 45,106 (32.1 %) | 13,600 (37.0 %) | 9,345 (35.1 %) | 6,540 (34.3 %) | 5,922 (31.3 %) | 7,176 (29.1 %) |
| PODI ≥ 2 | 8,884 (6.3 %) | 2,648 (7.2 %) | 1,891 (7.1 %) | 1,257 (6.6 %) | 1,135 (6.0 %) | 1,476 (6.0 %) |
| PCCC |  |  |  |  |  |  |
| PCCC = 0 | 31,380 (22.3 %) | 7,506 (20.4 %) | 5,938 (22.3 %) | 4,445 (23.3 %) | 4,318 (22.8 %) | 6,273 (25.4 %) |
| PCCC = 1 | 42,425 (31.2 %) | 11,052 (30.0 %) | 7,882 (29.6 %) | 5,681 (29.8 %) | 5,385 (28.4 %) | 7,168 (29.0 %) |
| PCCC ≥ 2 | 66,704 (47.5 %) | 18,242 (49.6 %) | 12,778 (48.0 %) | 8,918 (46.8 %) | 9,246 (48.8 %) | 11,239 (45.5 %) |
| ECMO, n (%) | 1,257 (0.89 %) | 403 (1.10 %) | 286 (1.08 %) | 191 (1.00 %) | 127 (0.72 %) | 181 (0.73 %) |
| Dialysis, n (%) | 1,581 (1.13 %) | 508 (1.38 %) | 351 (1.32 %) | 206 (1.08 %) | 210 (1.11 %) | 233 (0.94 %) |
| Surgery, n (%) | 71,761 (51.1 %) | 19,873 (54.0 %) | 14,205 (53.4 %) | 10,478 (55.0 %) | 10,238 (54.0 %) | 12,234 (49.6 %) |
| ICD chapters of main discharge diagnoses |  |  |  |  |  |  |
| Unknown | 74 (0.1 %) | 17 (0.1 %) | 18 (0.1 %) | 12 (0.1 %) | 9 (0.1 %) | 14 (0.1 %) |
| I: Infections | 5,988 (4.3 %) | 1,402 (3.8 %) | 1,002 (3.8 %) | 712 (3.7 %) | 713 (3.8 %) | 1087 (4.4 %) |
| II: Neoplasms | 10,933 (7.8 %) | 2,502 (6.8 %) | 1,726 (6.5 %) | 1,335 (7.0 %) | 1442 (7.6 %) | 1554 (6.3 %) |
| III: Blood | 1,954 (1.4 %) | 423 (1.2 %) | 357 (1.3 %) | 270 (1.4 %) | 237 (1.3 %) | 307 (1.2 %) |
| IV: Endocrine | 4,141 (3.0 %) | 1,111 (3.0 %) | 695 (2.6 %) | 601 (3.2 %) | 562 (3.0 %) | 804 (3.3 %) |
| IX: Circulatory | 8,419 (6.0 %) | 2,042 (5.6 %) | 1,579 (5.9 %) | 1,055 (5.5 %) | 1038 (5.5 %) | 1345 (5.5 %) |
| V: Mental | 1,918 (1.4 %) | 497 (1.4 %) | 396 (1.5 %) | 265 (1.4 %) | 218 (1.2 %) | 353 (1.4 %) |
| VI: Nervous | 9,363 (6.7 %) | 2,612 (7.1 %) | 1,823 (6.9 %) | 1,136 (6.0 %) | 1311 (6.9 %) | 1698 (6.9 %) |
| VII: Eye | 295 (0.2 %) | 60 (0.2 %) | 47 (0.2 %) | 45 (0.2 %) | 65 (0.3 %) | 43 (0.2 %) |
| VIII: Ear | 478 (0.3 %) | 86 (0.2 %) | 102 (0.4 %) | 64 (0.3 %) | 69 (0.4 %) | 77 (0.3 %) |
| X: Respiratory | 24,173 (17.2 %) | 6,675 (18.1 %) | 4,684 (17.6 %) | 3,103 (16.3 %) | 3048 (16.1 %) | 4265 (17.3 %) |
| XI: Digestive | 8,323 (5.9 %) | 2,090 (5.7 %) | 1,449 (5.5 %) | 1,129 (5.9 %) | 1280 (6.8 %) | 1623 (6.6 %) |
| XII: Skin | 416 (0.3 %) | 106 (0.3 %) | 81 (0.3 %) | 44 (0.2 %) | 53 (0.3 %) | 81 (0.3 %) |
| XIII: Musculoskeletal | 3,613 (2.6 %) | 941 (2.6 %) | 717 (2.7 %) | 560 (2.9 %) | 565 (3.0 %) | 681 (2.8 %) |
| XIV: Genitourinary | 2,114 (1.5 %) | 560 (1.5 %) | 425 (1.6 %) | 267 (1.4 %) | 253 (1.3 %) | 438 (1.8 %) |
| XIX: Injury-Poisoning | 19,933 (14.2 %) | 5,330 (14.5 %) | 4,006 (15.1 %) | 3,035 (15.9 %) | 2807 (14.8 %) | 3697 (15.0 %) |
| XV: Pregnancy | 97 (0.1 %) | 27 (0.1 %) | 16 (0.1 %) | 17 (0.1 %) | 8 (0.04 %) | 24 (0.1 %) |
| XVI: Perinatal | 503 (0.4 %) | 104 (0.3 %) | 102 (0.4 %) | 60 (0.3 %) | 96 (0.5 %) | 119 (0.5 %) |
| XVII: Congenital | 32,203 (22.9 %) | 8,942 (24.3 %) | 6,315 (23.7 %) | 4,606 (24.2 %) | 4438 (23.4 %) | 5403 (21.9 %) |
| XVIII: Symtopms - Findings | 4,032 (2.9 %) | 993 (2.7 %) | 773 (2.9 %) | 544 (2.9 %) | 480 (2.5 %) | 774 (3.1 %) |
| XX: Heath status factors | 1,169 (0.8 %) | 184 (0.5 %) | 213 (0.8 %) | 144 (0.8 %) | 207 (1.1 %) | 217 (0.9 %) |
| XXII: Codes for special purpose | 370 (0.3 %) | 96 (0.3 %) | 72 (0.3 %) | 40 (0.2 %) | 50 (0.3 %) | 76 (0.3 %) |

### Clinical results

Sex of CICT cases was female in 43.9 %, and mechanical ventilation was performed in 47,138 (33.5 %) cases (Table 1). A total of 3,386 (2.40 %) cases died, with the length of hospital stay similar between survivors (median 9 days (IQR 4 – 16)) and deceased cases (10 (3 – 33)). The PODI had 1 positive category in 32.1 % and ≥ 2 categories in 6.3 %. More than half (51.1 %) of cases received surgery during their hospital stay, while extracorporeal membrane oxygenation (ECMO) and dialysis were performed in 0.89 % and 1.13 %, respectively. One positive category of the PCCC was observed in 31.2 % and ≥ 2 categories in 47.5 %.

Notably, cases from more deprived quintiles had lower case fatality (2.35 % in Q5 vs. 2.64 % in Q1) along with lower organ dysfunction scores and fewer chronic conditions. Percentages of invasive measures such as mechanical ventilation, ECMO, and dialysis were also lower in higher deprivation with monotonous trends across GISD quintiles (Table 1).

### Incidence rates

IRs across GISD quintiles differed, with an almost linear increase of admissions with higher deprivation from 6,191/100,000 CY for the least deprived (Q1) up to 9,255/100,000 CY in the most deprived (Q5) (Figure 1). CICT IRs increased monotonously from 97/100,000 CY to 128/100,000 CY from Q1 to Q5. Mortality likewise increased from 2.54/100,000 CY to 2.96/100,000 CY (Figure 1). Over time, standardized incidence rates declined for admissions and CICT but not clearly for mortality at different rates depending on the GISD quintile (Table 2, figure 2).

**Figure 1:**
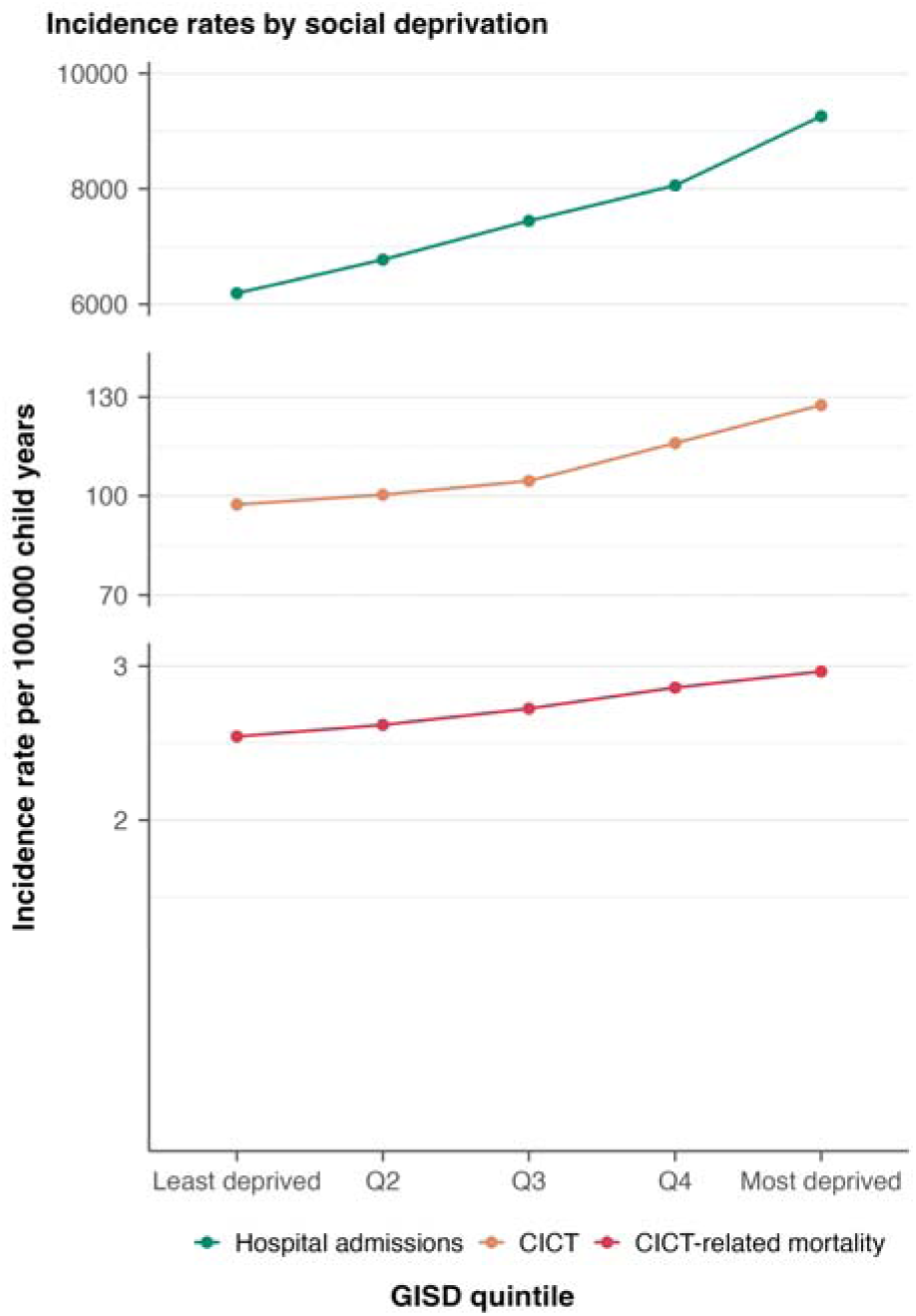
Incidence rates of hospital admissions, CICT, and CICT-related mortality by quintile of area-level social deprivation. CICT = complex intensive care treatment

**Figure 2:**
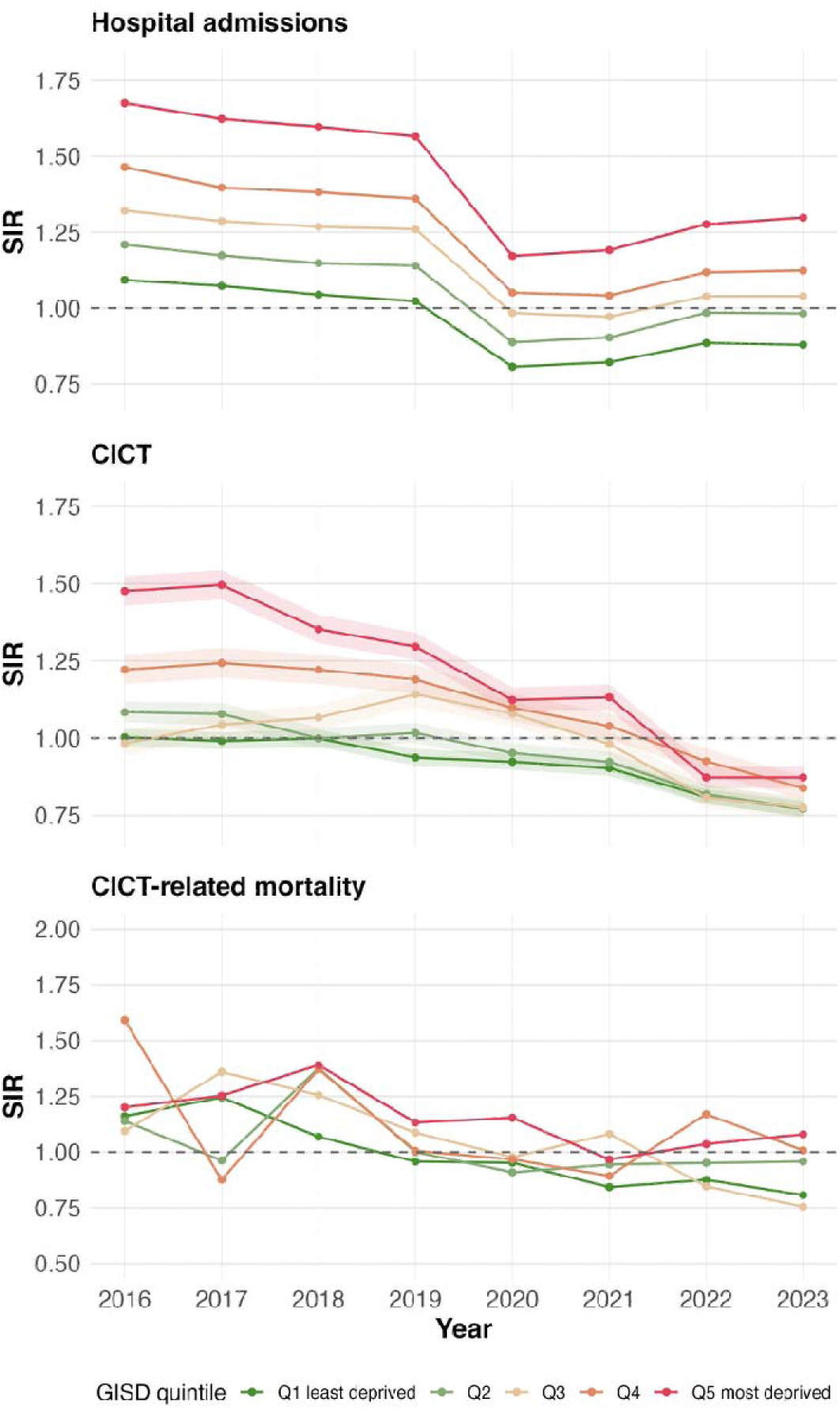
Standardized incidence rates of hospital admissions, CICT, and CICT-related mortality by quintile of area-level social deprivation over time. CICT = complex intensive care treatment, GISD = German index of social deprivation, SIR = standardized incidence rate.

**Table 2:**
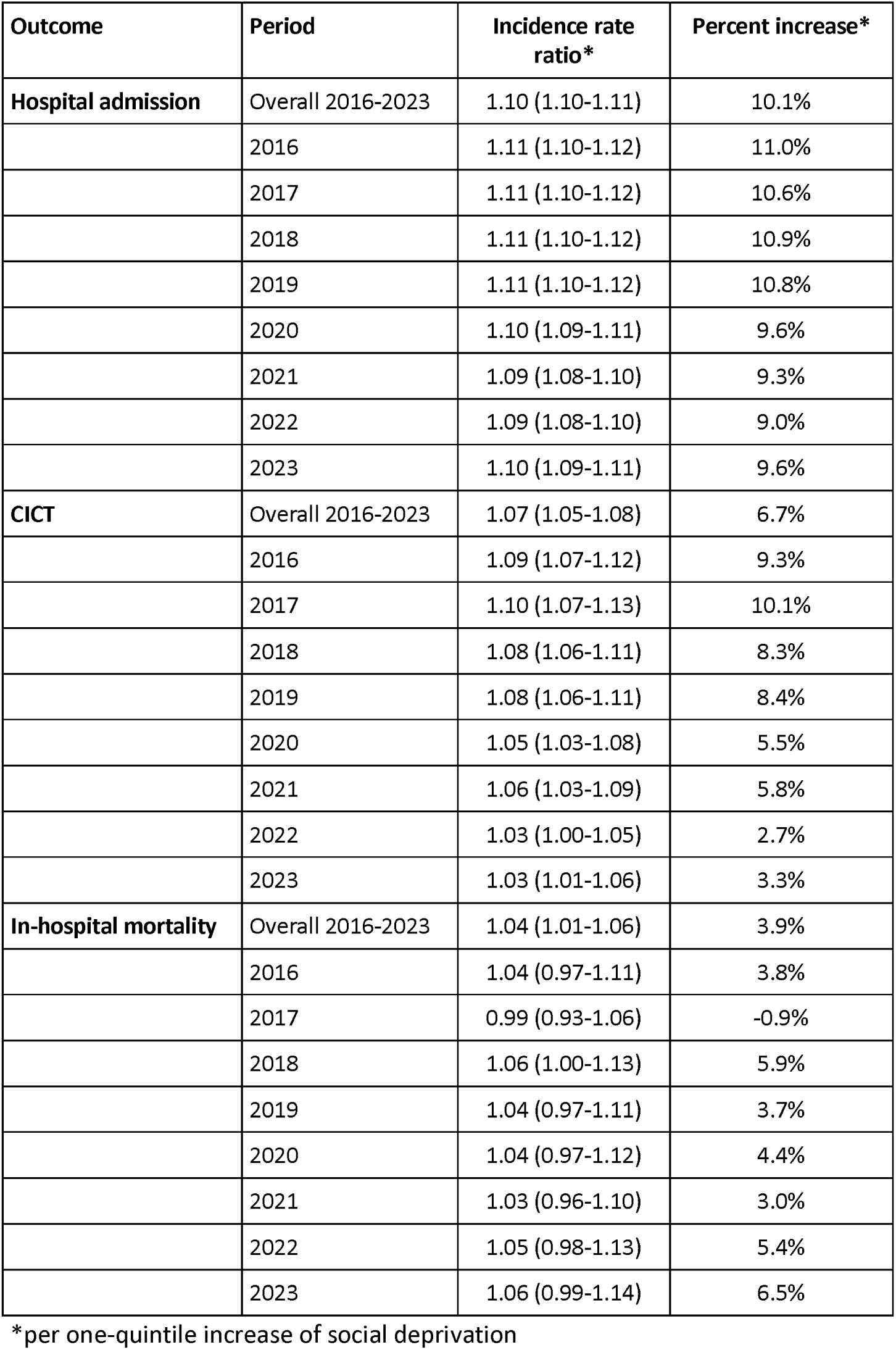
Population-based estimates of incidence rate ratio increase per quintile of social deprivation in children in Germany (2026 – 2023)

### Likelihood-ratio-test for model selection for IRR

LRT tests were significant for the admission and CICT models (p = 0.003 and 0.04). For mortality, the dispersion test was non-significant (p = 0.87), likely due to limited power given the small number of events. Given the pronounced, non-monotonic variability observed in the raw data, we used the negative binomial model nonetheless to avoid underestimating standard errors.

### Adjusted IRR

For each one-quintile increase of the GISD, the risk for admission, CICT, and mortality increased by 10.1 %, 6.7 %, and 3.9 %, respectively (adjusted IRRs: 1.10 (95 % CI 1.10-1.11), 1.07 (1.05-1.08), and 1.04 (1.01-1.06)). The interaction analysis showed that IRRs for admissions and CICT declined faster in the more deprived quintiles but remained stable for mortality (Table 2). Additional adjustment for age group did not materially alter these estimates (age-adjusted IRRs: 1.09 (95 % CI 1.08-1.11) for admission, 1.06 (1.04-1.08) for CICT, and 1.03 (1.00-1.06) for mortality), indicating that the deprivation gradient was not explained by differences in the age structure of the pediatric population across quintiles.

### Counterfactual scenario

Applying the IRs observed in the least deprived quintile (Q1) to the populations of Q2–Q5, we estimated 1,295,896 excess admissions (20.8 % of all admissions), 11,254 excess CICT cases (12.6 % of all CICT cases), and 194 excess deaths (8.7 % of all deaths) during the study period. The number of excess cases increased monotonously with increasing deprivation for all three outcomes (Table 3).

**Table 3:**
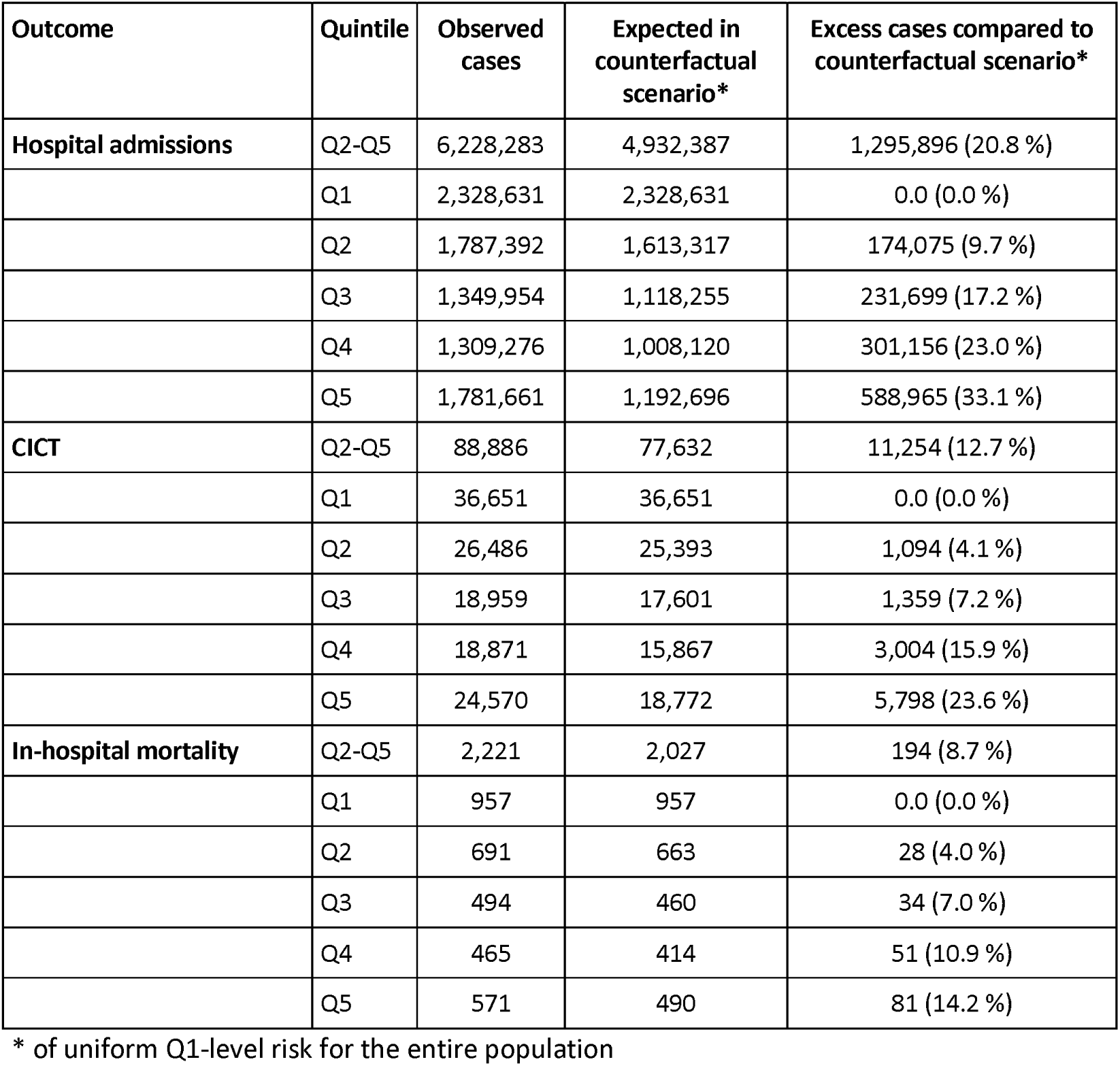
Excess cases assuming a best-case scenario (incidence rates as in least deprived cases for the entire population) by quintile of social deprivation.

The sensitivity analyses yielded consistent results pointing in the same overall direction. Differences were observed for the incidence of CICT in Q3 and Q4, which was slightly higher than in Q5 (Supplementary figure 2).

## Discussion

This nationwide, population-based analysis of pediatric hospital discharge data from Germany found a socioeconomic gradient in the population burden of hospital admission, CICT, and CICT-related mortality that persisted after adjustment for the age structure of the pediatric population. Although children from more deprived areas experienced higher population-level risks across all three outcomes, CICT cases from these areas had lower case fatality, fewer organ dysfunctions, and fewer chronic conditions, highlighting the distinction between population-level risk and case-level severity. While socioeconomic disparities in hospital admission and CICT narrowed over time, the mortality gradient remained unchanged, suggesting that progress in reducing inequalities has not been uniform across the continuum of pediatric care. These findings suggest that socioeconomic inequalities arise primarily before children reach the PICU, increasing the population burden of critical illness rather than worsening outcomes once intensive care has been initiated.

The observed socioeconomic gradient is consistent with previous studies reporting higher rates of hospital admission, pediatric intensive care use, and mortality among children from more deprived areas [2, 7–10]. To our knowledge, this is the first nationwide, full-census study outside the UK and North America to demonstrate these associations, indicating that universal health insurance alone does not eliminate socioeconomic disparities in pediatric critical illness. Previous German studies have reported inconsistent findings, ranging from little socioeconomic variation in pediatric hospital admissions [27] to higher inpatient healthcare utilization among children with lower socioeconomic status in the KiGGS Wave 2 study [28]. Our findings extend this evidence by demonstrating that the socioeconomic gradient also applies to severe illness requiring CICT and to population-level CICT-related mortality.

The apparent paradox of higher population-level incidence but lower case fatality among CICT cases from more deprived areas likely reflects differences in case-mix between quintiles rather than a protective effect of deprivation, highlighting the distinction between population-at-risk and case-fatality perspectives previously described in preterm birth statistics [29]. In less deprived areas, CICT cases may disproportionately comprise children with complex chronic conditions who survive to require intensive care partly because of better access to specialized outpatient and home-based care, whereas CICT cases in more deprived areas may more often arise from acute events with lower baseline severity. This interpretation aligns with earlier findings of a dual effect of deprivation on childhood hospital admissions, encompassing both organic pathology and impaired parental coping [3], and with evidence from the neonatal setting that mortality inequalities are only partly explained by case-mix at unit entry, with residual disparities attributable to other factors [6]. Residual inequalities may result from disparities in access to specialized care, as has been suggested for pediatric traumatic brain injury patients from areas with high deprivation, as they are more likely to be transferred [30].

While transportation to a specialized center is not a risk factor *per se* [31], a more concerning finding of our study are the divergent time trends across outcomes: the deprivation gradient in admissions and CICT narrowed over the study period but the gradient in mortality remained stable. This finding is particularly relevant because reductions in hospital utilization do not necessarily indicate that inequalities in the most severe forms of pediatric illness are improving. One possible explanation for the decline of hospital admissions is the ongoing restructuring of pediatric inpatient care in Germany. Even though pediatric ward capacity has declined substantially over the past decade [13, 14], the number of pediatric intensive care beds decreased only marginally [13].

Our findings emphasize that ICU outcomes alone only provide a partial picture of socioeconomic inequalities in pediatric critical illness. Although case fatality among children receiving CICT was lower in more deprived areas, the substantially higher incidence of CICT resulted in a markedly greater population burden of critical illness. These discrepancies translate into a relevant public health burden: under the counterfactual scenario of uniform least-deprived quintile incidences, > 1 million hospital admissions, > 10,000 CICT cases, and 194 CICT-related deaths were associated with the socioeconomic gradient. This aligns with findings from England, where a similarly steep deprivation gradient in child mortality suggested that over one-fifth of child deaths could be avoided if the most deprived half of the population experienced the same mortality as the least deprived [10]. Also in adults, lower socioeconomic status is associated with higher severity of disease at admission, length of ICU stays, and mortality [32, 33]. These findings may also have implications for regional planning of pediatric critical care resources, as areas with greater socioeconomic deprivation may generate a disproportionately higher demand for intensive care services across all ages.

While the ecological design of this study precludes causal inference on underlying mechanisms, unequal access to primary and preventive care, environmental exposures, and differences in social support structures in deprived areas represent plausible upstream contributors to the elevated population-level incidence of illness requiring hospital admission and CICT. As poverty and low socioeconomic status are passed on between generations, these findings point to potential prevention targets upstream in the care continuum, at the same time as underscoring the need for a better understanding of the mechanisms linking socioeconomic deprivation to pediatric critical illness.

This study has several limitations. Our definition of intensive care relied on CICT coding and therefore did not capture all ICU admissions. The transition in CICT coding rules after 2022, when some hospitals lost eligibility to code pediatric CICT, may have influenced observed time trends thereafter. As an area-level rather than individual-level measure, the GISD precludes conclusions about individual-level mechanisms and may mask within-municipality heterogeneity; similarly, updated GISD classifications were unavailable beyond 2021, so 2021 quintiles were also applied to 2022–2023 cases. Cases without a valid AGS were excluded from GISD-stratified analyses. We assumed that these were mainly from non-German residents – if missing AGS also occurred among German residents, incidence may have been slightly underestimated. Children who died before reaching the hospital are not represented in hospital discharge data at all, and children who died within the first 24 hours after admission generally do not fulfil the duration-based criteria for CICT coding and were therefore not counted as CICT cases. Both groups comprise the most severely ill children and this should be considered when interpreting the absolute numbers reported here. Finally, excess case estimates reflect the observed gradient under the counterfactual assumption of uniform Q1-level incidence and do not imply that this reduction is causally attributable to deprivation alone or readily achievable in practice.

## Conclusion

In this nationwide analysis of pediatric hospital discharge data from Germany, area-level socioeconomic deprivation was associated with a dose-dependent population-level incidence of hospital admission, CICT, and CICT-related mortality, despite Germany’s universal health insurance coverage. Notably, this population-level gradient coexisted with lower case fatality and illness severity among CICT cases from more deprived areas, underscoring the importance of distinguishing between population-level mortality and case fatality when interpreting socioeconomic disparities. The stability of the mortality gradient over time, in contrast to a narrowing gradient in admissions and CICT, suggests that progress in reducing socioeconomic disparities in pediatric critical illness has been uneven across the care pathway. These findings call for further research into the modifiable drivers of these disparities.

## Supporting information

RECORD Checklist

## Data Availability

The original dataset remains at the Federal Statistical Office and can be accessed by qualified researchers at designated research data centers after filing a request and signing a confidentiality agreement. The data generated for this study and exported from the Federal Statistical Office will be made available upon reasonable request.

## List of abbreviations

AGS: Amtlicher Gemeindeschlüssel (official German municipality code)
aIRR: Adjusted incidence rate ratio
CI: Confidence interval
CICT: Complex intensive care treatment
CY: Child years
ECMO: Extracorporeal membrane oxygenation
GHD: German hospital discharge data
GISD: German Index of Socioeconomic Deprivation
ICD-10: International Statistical Classification of Diseases and Related Health Problems, 10th revision
ICU: Intensive care unit
IQR: Interquartile range
IR: Incidence rate
IRR: Incidence rate ratio
LRT: Likelihood-ratio test
OPS: Operationen- und Prozedurenschlüssel (German Operation and Procedure System)
PCCC: Pediatric Complex Chronic Conditions system
PICU: Pediatric intensive care unit
PODI: Pediatric Organ Dysfunction Index
pSOFA: Pediatric Sequential Organ Failure Assessment SD Standard deviation
SIR: Standardized incidence ratio

## Statements and Declarations

### Ethics approval and consent to participate

Only secondary fully anonymized data were used that did not require an ethics approval.

### Consent for publication

N/A

### Authors’ contributions

Study design: NB; data extraction and analyses: RH and NB; visualization: NB; drafting initial manuscript: NB; interpretation of study results and revision of manuscript draft: UFM, CDS, CO, HR, RH, CH.

## Acknowledgements

None.

