## Supplementary materials for "Socioeconomic deprivation and time trends in pediatric hospital admissions, intensive care treatment, and mortality: a nationwide population-based study in Germany"

**Supplementary table 1:** Patient level and structural inclusion criteria for the application of the operation and procedure code for complex intensive care treatment

|  | **Pediatric CICT** | **Adult CICT** |
| --- | --- | --- |
| Operation and procedure code | 8-98d | 8-980, 8-98f |
| Age | Patients ≥ 28 days old (and ≥ 2,500g) up to the age of 18 years at admission | Patients ≥ 18 years (or from 14 years in specific settings) |
| Medical Leadership | Specialist in Pediatrics (or Pediatric Surgery) with the additional qualification "Pediatric Intensive Care" | Specialist in any field with the additional qualification "Intensive Care Medicine" |
| ICU type | Specifically a pediatric intensive care unit  (since 2021, with transitional phase in 2021/2022) | Intensive care unit |
| Medical Presence | Constant 24h presence of a **pediatric specialist** with a sub-specialization in intensive care | Constant 24h presence of a **specialist** (e.g., Anesthesia, Internal Medicine, Surgery) with a sub-specialization in intensive care |
| Exclusion Criteria | Mere "intensive monitoring" without acute treatment of vital organ systems | |
| Short-term Care | Excludes short-term (< 24h) stabilization after surgery. | |

CICT = complex intensive care treatment, ICU = intensive care unit

**Supplementary figures**

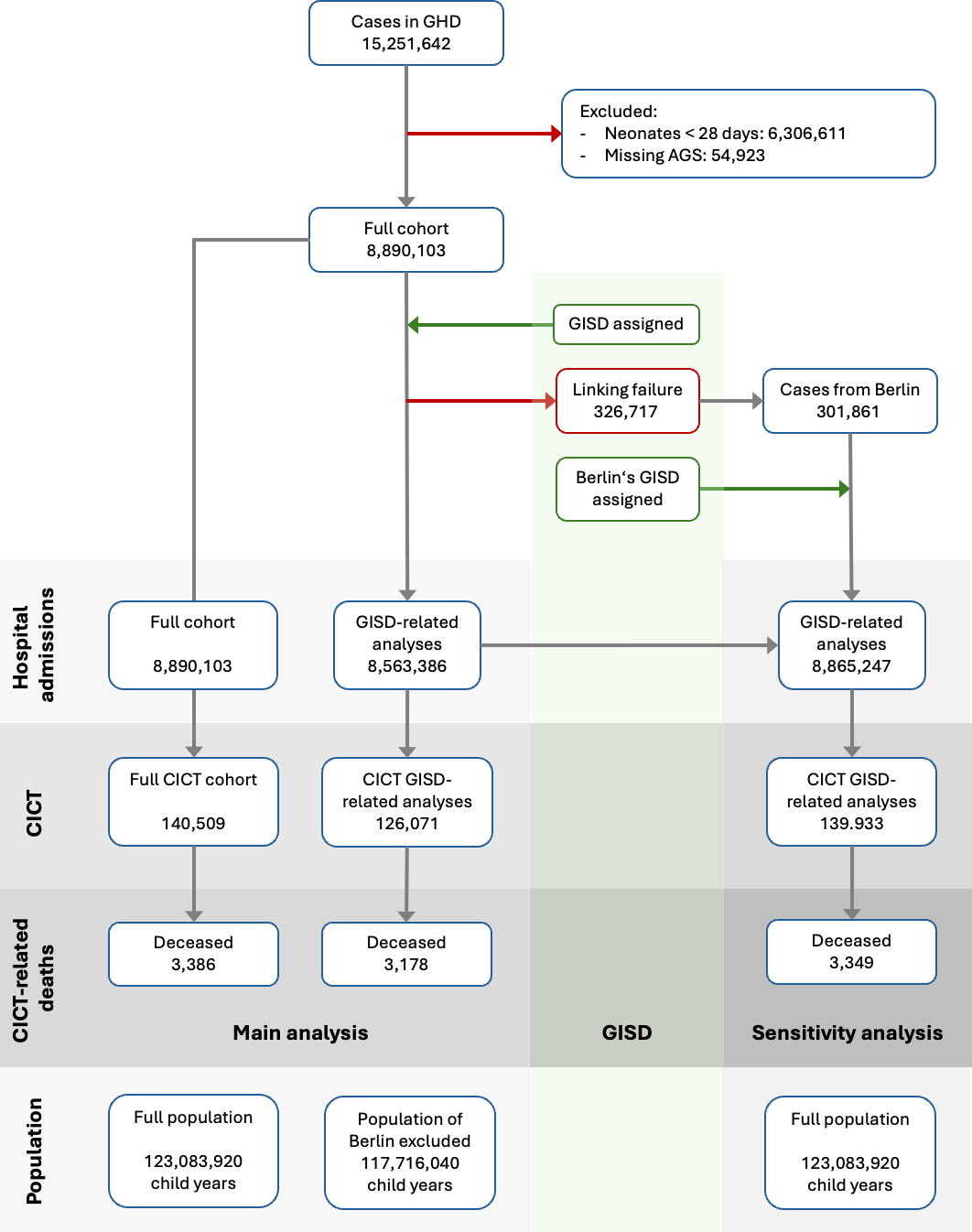

**Supplementary figure 1:** Flow chart of analyzed cases.

AGS = official municipality code, CICT = complex intensive care treatment, GHD = German hospital dataset, GISD = German index of social deprivation

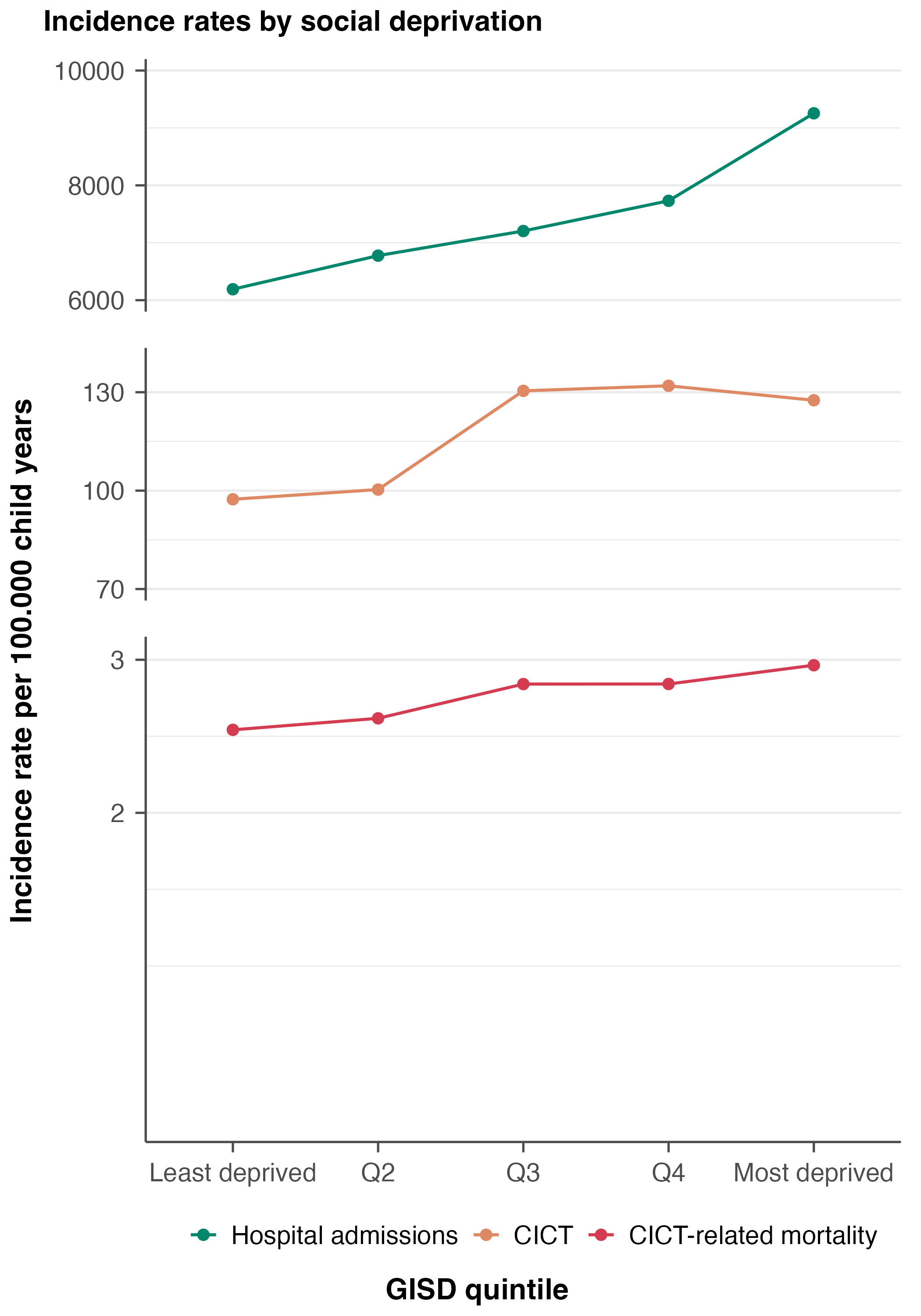

**Supplementary figure 2:** Sensitivity analysis for incidence rates of hospital admissions, CICT, and CICT-related mortality by quintile of area-level social deprivation with Berlin cases assigned the respective years’ quintiles.

CICT = complex intensive care treatment
