## Supplementary material for "Socioeconomic deprivation and time trends in pediatric hospital admissions, intensive care treatment, and mortality: a nationwide population-based study in Germany": RECORD Checklist

**The RECORD statement – checklist of items, extended from the STROBE statement, that should be reported in observational studies using routinely collected health data.**

|  | **Item No.** | **STROBE items** | **Location in manuscript where items are reported** | **RECORD items** | **Location in manuscript where items are reported** |
| --- | --- | --- | --- | --- | --- |
| **Title and abstract** | | | | | |
|  | 1 | (a) Indicate the study’s design with a commonly used term in the title or the abstract (b) Provide in the abstract an informative and balanced summary of what was done and what was found | Title p. 1; Abstract p. 2 | RECORD 1.1: The type of data used should be specified in the title or abstract. When possible, the name of the databases used should be included.  RECORD 1.2: If applicable, the geographic region and timeframe within which the study took place should be reported in the title or abstract.  RECORD 1.3: If linkage between databases was conducted for the study, this should be clearly stated in the title or abstract. | 1.1: p. 1 (title: nationwide population-based study, Germany); p. 2 (Abstract – Methods: complete German hospital discharge data, GISD).  1.2: p. 1 (title: Germany); p. 2 (Abstract – Methods: 2016 to 2023).  1.3: p. 2 (Abstract – Methods: cases linked to the GISD via municipality of residence) |
| **Introduction** | | | | | |
| Background rationale | 2 | Explain the scientific background and rationale for the investigation being reported | p. 3 (Introduction) |  |  |
| Objectives | 3 | State specific objectives, including any prespecified hypotheses | p. 3 (Introduction, last paragraph: aim of the study) |  |  |
| **Methods** | | | | | |
| Study Design | 4 | Present key elements of study design early in the paper | p. 2 (Abstract – Methods); p. 3 (Methods, first paragraph) |  |  |
| Setting | 5 | Describe the setting, locations, and relevant dates, including periods of recruitment, exposure, follow-up, and data collection | p. 3–4 (Methods – Data source; Case selection); p. 4 (Determination of socioeconomic deprivation; Population at risk) |  |  |
| Participants | 6 | *(a) Cohort study* - Give the eligibility criteria, and the sources and methods of selection of participants. Describe methods of follow-up  *Case-control study* - Give the eligibility criteria, and the sources and methods of case ascertainment and control selection. Give the rationale for the choice of cases and controls  *Cross-sectional study* - Give the eligibility criteria, and the sources and methods of selection of participants  *(b) Cohort study* - For matched studies, give matching criteria and number of exposed and unexposed  *Case-control study* - For matched studies, give matching criteria and the number of controls per case | p. 4 (Methods – Case selection; Complex intensive care treatment); p. 6 (Results – Overall numbers and matching); Supplementary figure 1 (Supplementary material, p. 2) | RECORD 6.1: The methods of study population selection (such as codes or algorithms used to identify subjects) should be listed in detail. If this is not possible, an explanation should be provided.  RECORD 6.2: Any validation studies of the codes or algorithms used to select the population should be referenced. If validation was conducted for this study and not published elsewhere, detailed methods and results should be provided.  RECORD 6.3: If the study involved linkage of databases, consider use of a flow diagram or other graphical display to demonstrate the data linkage process, including the number of individuals with linked data at each stage. | 6.1: p. 4 (Case selection; Complex intensive care treatment); criteria and codes for CICT in Supplementary table 1 (Supplementary material, p. 1)  6.2: p. 4 (GISD validation, refs [15, 24]); PODI ref [25]; PCCC ref [26], applied with modifications per ref [12]  6.3: Supplementary figure 1 (Supplementary material, p. 2; flow chart with numbers per selection and linkage step); p. 6 (Results – Overall numbers and matching) |
| Variables | 7 | Clearly define all outcomes, exposures, predictors, potential confounders, and effect modifiers. Give diagnostic criteria, if applicable. | p. 4 (Methods – Complex intensive care treatment; Determination of socioeconomic deprivation; Organ dysfunction; Chronic conditions); p. 5 (Primary and secondary outcomes) | RECORD 7.1: A complete list of codes and algorithms used to classify exposures, outcomes, confounders, and effect modifiers should be provided. If these cannot be reported, an explanation should be provided. | 7.1: p. 4 and Supplementary table 1 (Supplementary material, p. 1; OPS-based criteria for CICT). Complete code lists for PODI and PCCC are not reproduced; the published methods are referenced (PODI [25], PCCC [26], modifications [12]). |
| Data sources/ measurement | 8 | For each variable of interest, give sources of data and details of methods of assessment (measurement).  Describe comparability of assessment methods if there is more than one group | p. 3–4 (Methods – Data source; Case selection; Determination of socioeconomic deprivation; Population at risk); p. 4 (Organ dysfunction; Chronic conditions) |  |  |
| Bias | 9 | Describe any efforts to address potential sources of bias | p. 5 (Methods – Missing data, incl. missing AGS and linkage failure; cell-size censoring); p. 6 (sensitivity analyses); p. 9 (Discussion – limitations) |  |  |
| Study size | 10 | Explain how the study size was arrived at | p. 3–4 (complete nationwide census; no sample size calculation); p. 6 (Results – Overall numbers and matching) |  |  |
| Quantitative variables | 11 | Explain how quantitative variables were handled in the analyses. If applicable, describe which groupings were chosen, and why | p. 4 (GISD quintiles); p. 5 (Methods – Statistical analyses) |  |  |
| Statistical methods | 12 | (a) Describe all statistical methods, including those used to control for confounding  (b) Describe any methods used to examine subgroups and interactions  (c) Explain how missing data were addressed  (d) *Cohort study* - If applicable, explain how loss to follow-up was addressed  *Case-control study* - If applicable, explain how matching of cases and controls was addressed  *Cross-sectional study* - If applicable, describe analytical methods taking account of sampling strategy  (e) Describe any sensitivity analyses | p. 5–6 (Methods – Statistical analyses: descriptive statistics, incidence rates, negative binomial regression with population offset, likelihood-ratio tests, counterfactual estimation, sensitivity analyses incl. age adjustment); missing data p. 5 |  |  |
| Data access and cleaning methods |  | .. |  | RECORD 12.1: Authors should describe the extent to which the investigators had access to the database population used to create the study population.  RECORD 12.2: Authors should provide information on the data cleaning methods used in the study. | 12.1: p. 3 (Data source: complete §21 KHEntgG census; access to the dataset at the regional research data centre); p. 6 (Software: data extraction and analysis on site at the research data centre) 12.2: p. 5 (Methods – Missing data: age, diagnosis and department codes complete, OPS coding, missing AGS, linkage failure); p. 5 (statutory cell-size censoring, n = 43 deaths) |
| Linkage |  | .. |  | RECORD 12.3: State whether the study included person-level, institutional-level, or other data linkage across two or more databases. The methods of linkage and methods of linkage quality evaluation should be provided. | 12.3: p. 4 (area-level, not person-level linkage: GISD assigned at municipality level via the official municipality code (AGS); population denominators linked via AGS); p. 5 (Missing data – linkage failure, 204 municipality-free areas, Berlin handling and sensitivity analysis); p. 6 (Results – matching quality in clinical and population data) |
| **Results** | | | | | |
| Participants | 13 | (a) Report the numbers of individuals at each stage of the study (*e.g.*, numbers potentially eligible, examined for eligibility, confirmed eligible, included in the study, completing follow-up, and analysed)  (b) Give reasons for non-participation at each stage.  (c) Consider use of a flow diagram | p. 6 (Results – Overall numbers and matching in the clinical dataset / in the population data); Supplementary figure 1 (Supplementary material, p. 2) | RECORD 13.1: Describe in detail the selection of the persons included in the study (*i.e.,* study population selection) including filtering based on data quality, data availability and linkage. The selection of included persons can be described in the text and/or by means of the study flow diagram. | 13.1: p. 6 (Results – Overall numbers and matching: exclusions by age and missing AGS, unmatched cases and Berlin handling); Supplementary figure 1 (Supplementary material, p. 2; flow chart per step and analysis) |
| Descriptive data | 14 | (a) Give characteristics of study participants (*e.g.*, demographic, clinical, social) and information on exposures and potential confounders  (b) Indicate the number of participants with missing data for each variable of interest  (c) *Cohort study* - summarise follow-up time (*e.g.*, average and total amount) | p. 6–7 (Results – Clinical results); Table 1 (Tables document, p. 1–3) |  |  |
| Outcome data | 15 | *Cohort study* - Report numbers of outcome events or summary measures over time  *Case-control study* - Report numbers in each exposure category, or summary measures of exposure  *Cross-sectional study* - Report numbers of outcome events or summary measures | p. 6–7 (Results – Clinical results; Incidence rates); Table 1 (Tables, p. 1–3), Table 2 (Tables, p. 4); Figure 1, p. 15 |  |  |
| Main results | 16 | (a) Give unadjusted estimates and, if applicable, confounder-adjusted estimates and their precision (e.g., 95% confidence interval). Make clear which confounders were adjusted for and why they were included  (b) Report category boundaries when continuous variables were categorized  (c) If relevant, consider translating estimates of relative risk into absolute risk for a meaningful time period | p. 7 (Results – Incidence rates; Likelihood-ratio test; Adjusted IRR; Counterfactual scenario); Table 2 (Tables, p. 4), Table 3 (Tables, p. 5); Figure 1, p. 15 |  |  |
| Other analyses | 17 | Report other analyses done—e.g., analyses of subgroups and interactions, and sensitivity analyses | p. 7 (Results – sensitivity analyses; time trends); p. 6 (Methods – sensitivity analyses); Figure 2, p. 16; Supplementary figure 2 (Supplementary material, p. 3) |  |  |
| **Discussion** | | | | | |
| Key results | 18 | Summarise key results with reference to study objectives | p. 7 (Discussion, first paragraph) |  |  |
| Limitations | 19 | Discuss limitations of the study, taking into account sources of potential bias or imprecision. Discuss both direction and magnitude of any potential bias | p. 9 (Discussion – limitations) | RECORD 19.1: Discuss the implications of using data that were not created or collected to answer the specific research question(s). Include discussion of misclassification bias, unmeasured confounding, missing data, and changing eligibility over time, as they pertain to the study being reported. | 19.1: p. 9 (Discussion – limitations); p. 5 (Methods – Missing data) |
| Interpretation | 20 | Give a cautious overall interpretation of results considering objectives, limitations, multiplicity of analyses, results from similar studies, and other relevant evidence | p. 7–9 (Discussion) |  |  |
| Generalisability | 21 | Discuss the generalisability (external validity) of the study results | p. 8 (Discussion); p. 9 (Discussion – limitations) |  |  |
| **Other Information** | | | | | |
| Funding | 22 | Give the source of funding and the role of the funders for the present study and, if applicable, for the original study on which the present article is based | p. 11 (Statements and Declarations – Funding: none) |  |  |
| Accessibility of protocol, raw data, and programming code |  | .. |  | RECORD 22.1: Authors should provide information on how to access any supplemental information such as the study protocol, raw data, or programming code. | 22.1: p. 11 (Availability of data and materials); p. 6 (Software). No study protocol or programming code is currently made available. |

*Reference: Benchimol EI, Smeeth L, Guttmann A, Harron K, Moher D, Petersen I, Sørensen HT, von Elm E, Langan SM, the RECORD Working Committee. The REporting of studies Conducted using Observational Routinely-collected health Data (RECORD) Statement. *PLoS Medicine* 2015; in press.

*Checklist is protected under Creative Commons Attribution ([CC BY](http://creativecommons.org/licenses/by/4.0/)) license.
